# Multi-omics of stressful life events

**DOI:** 10.64898/2026.05.22.26353900

**Authors:** Elissar Azzi, Aino Heikkinen, Gabin Drouard, Teemu Palviainen, Colette S. Kabrita, Jaakko Kaprio, Miina Ollikainen

## Abstract

Stressful life events (SLEs) are associated with increased risk of psychiatric and somatic disease, yet the molecular correlates of stress exposure across time remain incompletely characterised. We conducted a multi-omic analysis in the Finnish Twin Cohort, examining genomic (*n* = 8,286), epigenomic (*n* = 387), proteomic (*n* = 401) and metabolomic (*n* = 434) data across three exposure windows: recent (≤6 months), proximal (≤5 years) and lifetime. Genome-wide association analysis identified a single significant locus on chromosome 1 (lead SNP rs10158287, *p* = 9.7 × 10^−9^), mapping to *DAB1*/*C8B*, with immune cell-specific eQTL effects in Th1/Th17 CD4^+^ T cells and cross-trait links to cardiometabolic risk; common variants explained 33.6% of variance in SLE scores; the *DAB1*/*C8B* locus alone accounted for 0.4%, consistent with a highly polygenic architecture. No epigenome-wide significant associations or relationships with epigenetic ageing measures (PCPhenoAge, DunedinPACE, PCGrimAge) were detected. Circulating molecular layers showed temporally structured signatures. Recent SLEs were characterised by coordinated immune-metabolic changes, including five proteins (IL-1β, TIGIT, Nectin-1, Carnosinase-1, Calcyphosin; all decreased) and 16 metabolites predominantly reflecting reduced HDL-related lipids and broader lipid pools, alongside enrichment of the lipoprotein assembly and clearance pathway and lower thyroid and lung proteomic age estimates. Proximal exposure showed no significant single-analyte associations but convergent negative pathway-level signals involving cell-cycle and centrosome-associated processes. Lifetime SLEs were characterised by immune-vascular and tissue-remodelling signatures, including ten proteins (all increased), enrichment of TNF/IL-10 signalling, cellular maintenance and epithelial differentiation pathways, and higher arterial proteomic age. These findings indicate that molecular correlates of stress exposure are temporally contingent rather than uniformly accumulating across the life course, with convergent involvement of adaptive immune regulation alongside metabolic and vascular remodelling.

## Introduction

Stressful life events (SLEs), including experiences such as bereavement, relationship disruptions, financial hardship and serious illness, constitute a major source of psychosocial adversity across the life course. Epidemiological studies have consistently linked exposure to such events with increased risk of a wide array of adverse health outcomes, including mental disorders, infectious disease susceptibility, cardiovascular and metabolic diseases, cancer and premature mortality.^1,2,3,4,5^ For example, spousal bereavement is associated with sharply elevated short-term mortality, particularly from cardiovascular causes,^6^ marital separation has repeatedly been linked to increased mortality,^7^ and employment loss is associated with adverse health outcomes, with disease risk varying by length of follow-up.^5^ These findings suggest that SLEs act through shared biological mechanisms relevant to multiple disease outcomes. The associations are thought to arise through repeated or sustained engagement of stress-regulatory processes that integrate endocrine, immune and metabolic responses, which over time can generate cumulative physiological wear-and-tear, often conceptualised as allostatic load.^8^

Although SLEs are often considered environmental exposures, evidence from twin and family studies indicates that inter-individual differences in exposure to and reporting of SLEs are partly shaped by genetic factors, with liability to certain categories of life stress displaying modest to moderate heritability (∼27–47%).^9^ At the molecular level, a growing body of evidence indicates that exposure to SLEs may become molecularly embedded through coordinated alterations across multiple regulatory layers. Epigenetic studies of psychosocial adversity, particularly in early-life exposure, support links to immune-inflammatory regulation and, in some contexts, faster epigenetic ageing.^10,11^ SLEs have additionally been associated with increased breast cancer risk, with DNA methylation changes implicated as a potential mechanistic link between stress exposure and disease risk.^4^ Proteomic studies in related stress-exposed contexts have demonstrated that recent trauma is associated with adaptive homeostasis pathway activation, whereas chronic post-traumatic stress disorder (PTSD) is characterised by signatures of inflammation, neurodegeneration and cardiometabolic dysfunction.^12^ Metabolomic analyses have reported stress-related perturbations in lipid and amino acid metabolism.^12,13^

Despite this literature, multi-omics investigations of common SLEs in population-based samples remain limited. Most prior work has focused on a single molecular layer or on extreme forms of adversity such as childhood maltreatment or PTSD, leaving uncertainty about how the timing and accumulation of stress shape molecular profiles across regulatory systems. To address this gap, we leverage the Finnish Twin Cohort, a longitudinal population-based cohort with detailed SLE assessments and linked genome-wide genotype, DNA methylation array, plasma proteomic and ^1^H-NMR metabolomic data. By distinguishing between recent (≤6 months), proximal (≤5 years) and lifetime SLE exposure, we aimed to identify molecular signatures reflecting both transient and more stable stress-related processes and to determine whether molecular correlates accumulate linearly or shift qualitatively across time.

## Materials and Methods

### Study population

The Older Finnish Twin Cohort consists of same-sex twin pairs born before 1958, recruited via a baseline postal survey in 1975 and followed up with comprehensive health and lifestyle questionnaires in 1981, 1990 and 2011.^14^ The life event inventory was included in both the 1981 and 2011 surveys. A total of 24 684 and 8 410 participants returned complete answers to the 1981 and 2011 surveys, respectively; the 2011 wave was restricted to participants born 1945–1957. Genome-wide association analyses used the 1981 SLE assessment, reflecting the larger sample available for genetic analyses. Given the time-invariant nature of germline variation, use of this earlier assessment does not constrain genotype–phenotype inference. Epigenomic, proteomic and metabolomic analyses used SLE assessments from the 2011 questionnaire, aligned with biospecimen collection from the EH-Epi in-person study (2012–2014), during which fasting blood samples were collected.^15^ Participants underwent measurement of blood pressure, height, weight, hip and waist circumference and completed interviews and questionnaires capturing health status and lifestyle alongside collection of blood samples used for the omics analysis. At the time of sampling, participants were aged 56–70 years (mean 62 years, SD ∼3 years; mean time since 2011 survey: 2.01 years, SD 0.7 years). Sample sizes for each omic layer are detailed in Table 1.

**Table 1.**
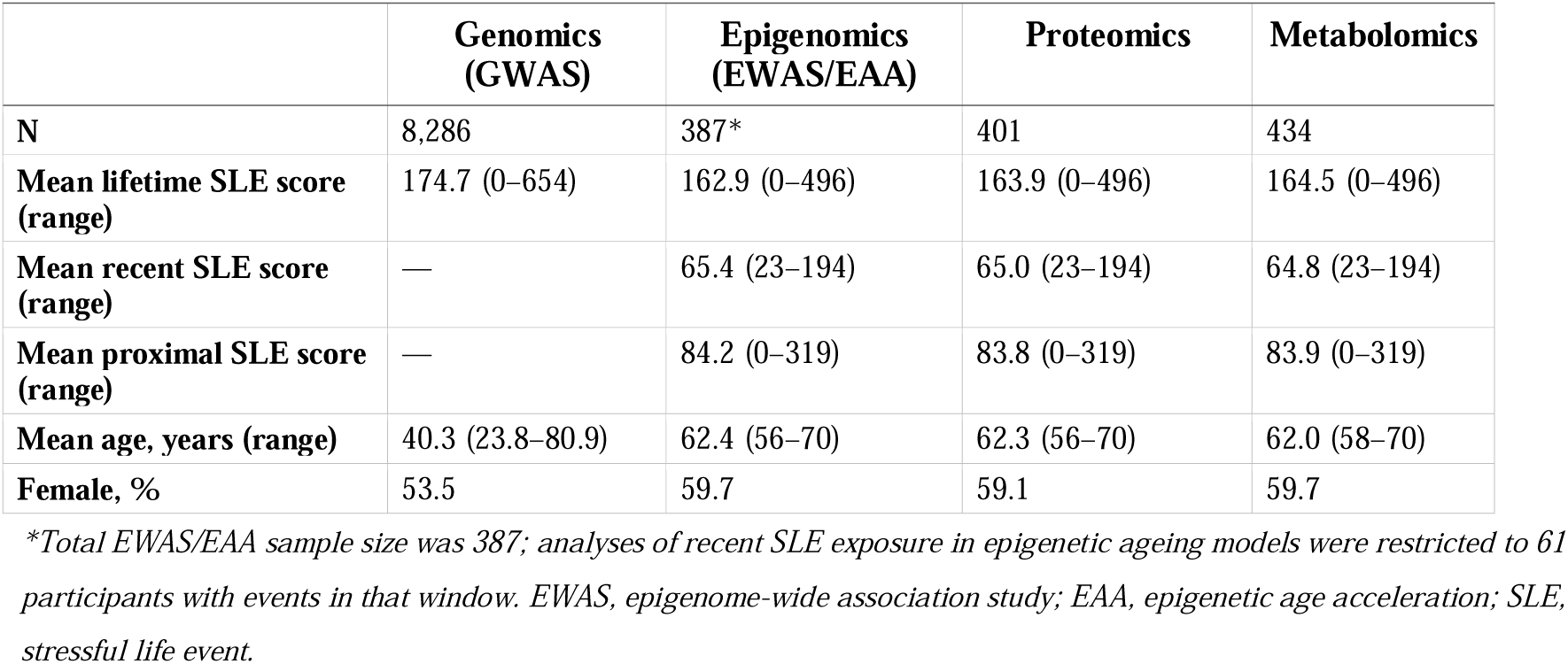
Cohort characteristics by molecular layer.

|  | **Genomics**<br>**(GWAS)** | **Epigenomics**<br>**(EWAS/EAA)** | **Proteomics** | **Metabolomics** |
| --- | --- | --- | --- | --- |
| **N** | 8,286 | 387* | 401 | 434 |
| **Mean lifetime SLE score**<br>**(range)** | 174.7 (0–654) | 162.9 (0–496) | 163.9 (0–496) | 164.5 (0–496) |
| **Mean recent SLE score**<br>**(range)** | — | 65.4 (23–194) | 65.0 (23–194) | 64.8 (23–194) |
| **Mean proximal SLE score**<br>**(range)** | — | 84.2 (0–319) | 83.8 (0–319) | 83.9 (0–319) |
| **Mean age, years (range)** | 40.3 (23.8–80.9) | 62.4 (56–70) | 62.3 (56–70) | 62.0 (58–70) |
| **Female, %** | 53.5 | 59.7 | 59.1 | 59.7 |

\*Total EWAS/EAA sample size was 387; analyses of recent SLE exposure in epigenetic ageing models were restricted to 61 participants with events in that window. EWAS, epigenome-wide association study; EAA, epigenetic age acceleration; SLE, stressful life event.

### Stressful life event assessment

SLEs were assessed using a life event inventory in which participants indicated which events they had experienced and the time elapsed since each event. The inventory encompasses items corresponding to the Holmes and Rahe Social Readjustment Rating Scale,^1,16^ capturing a broad range of life experiences across three time windows: “during the previous 6 months” (recent), “during the previous 5 years but not the past 6 months” (proximal), and “earlier in life” (lifetime/historic) and “never”. Each event was assigned a standardised stress weight (life change units) reflecting its relative severity, including: death of a spouse (100), death of a child (100), death of a close relative or friend (50), change in health status of a family member (44), difficulties in sexual life (39), significant work-related difficulties (23), substantial financial worsening (38), divorce or separation (69), breakdown of a long-term relationship (37), loss of employment (47), marked increase in marital conflicts (35) and illness or injury causing >3 weeks work incapacity (53). An SLE score was computed by summing the weights of all endorsed events within each exposure window. Exposures were classified as: recent (any event occurring within 6 months preceding the survey), proximal (events within the preceding 5 years), and lifetime (any event reported to have occurred at any point in life).

### Genotype data processing

DNA was extracted from blood or saliva samples collected between 1994 and 2018. Genotyping was performed using Illumina Human610-Quad, Human670-QuadCustom, HumanCoreExome 12 v1.0 A, 12 v1.1 A, 24 v1.0 A, 24 v1.1 A, 24 v1.2 A (GenCall) and Affymetrix FinnGen Axiom arrays (AxiomGT1), with genotype quality control performed in three batches.^17^ Pre-phasing used Eagle v2.3^18^ and imputation used Minimac3 v2.0.1 via the University of Michigan Imputation Server,^19^ with the Haplotype Reference Consortium release 1.1 reference panel.^20^ Post-imputation quality control excluded variants with effect-allele frequency <1%, Hardy–Weinberg equilibrium *p* < 1 × 10^−6^, or imputation INFO score <0.70.

### Epigenetic data processing

High-molecular-weight DNA was extracted from peripheral venous blood samples collected between 2011 and 2014 and bisulphite-converted using the EZ-96 DNA/Methylation-Gold Kit (Zymo Research). DNA methylation was quantified using Illumina Infinium HumanMethylation450 arrays. Preprocessing used the *meffil* R package,^21^; the full preprocessing pipeline is described in Hukkanen et al. (2025) including exclusion of low-quality samples (median methylated signal >3 SD, control probe values >5 SD, detection *p* > 0.01 or bead count <3), removal of low-quality probes (detection *p* > 0.01 or bead count <3 in >20% of samples), and exclusion of sex chromosome and cross-reactive probes.^22,23^ Between-sample quantile normalisation was applied, followed by within-sample Beta Mixture Quantile Normalisation (BMIQ) to adjust for type II probe bias.^24^ Epigenetic clocks were calculated using principal component-based (PC) implementations of GrimAge and PhenoAge,^25,26^ and DunedinPACE was computed to capture the pace of biological ageing.^27^ Epigenetic age acceleration (EAA) was defined as the residual from regressing estimated epigenetic age on chronological age. Associations between EAA measures and SLEs were examined using GEE models, with EAA measures specified as outcomes and lifetime or proximal SLE scores as predictors. Models were run unadjusted (Model 1) and adjusted for BMI, age, sex and smoking status (Model 2). p-values < 0.05 were considered statistically significant.

### Proteomic data processing

Proteomic data were generated from blood plasma samples analysed by proximity extension assay (Olink Explore 3072; Olink Proteomics AB, Uppsala, Sweden), quantifying 2 321 plasma proteins across cardiometabolic, inflammation, neurology and oncology panels following published preprocessing procedures.^28^ Briefly, outlier samples and assays failing Olink internal quality control criteria were excluded; proteins with normalised protein expression (NPX) values above the plate-specific limit of detection in ≥80% of samples were retained; and values below the limit of detection (<1% of data points) were replaced with the plate-specific limit of detection value. Following data processing, a total of 2,321 plasma proteins were available for 401 twins. Proteomic age clocks were calculated following published procedures for 22 systemic and organ-specific clocks, trained to predict chronological age, mortality or healthspan in UK Biobank participants, including the Proteomic Aging Clock (PAC), ProtAge, Conventional, and Healthspan Proteomic Score, as well as organ-specific clocks derived by Goeminne et al.^29,30,31,32^

### Metabolomic data processing

Metabolomic data were generated from the same blood samples using high-throughput proton nuclear magnetic resonance (^1^H-NMR) spectroscopy (Nightingale Health Ltd., Helsinki, Finland). Preprocessing followed published procedures.^28^ Metabolite values below the limit of quantification were coded as missing, as were values deviating >5 SD from the sample mean. Metabolites with >10% missingness were excluded; remaining missing values were imputed using the sample minimum per metabolite. Outliers were assessed by examining the first three principal components. An inverse normal rank transformation was applied to each metabolite to approximate Gaussian distributions.

### Statistical analyses

Genome-wide association analyses of lifetime SLE scores were conducted in 8 286 individuals using mixed-effects linear models implemented in GCTA-fastGWA,^33^ with age and sex as fixed covariates and a sparse genetic relationship matrix as a random effect. The genome-wide significance threshold was *p* < 5 × 10^-8^. Bayesian fine-mapping was performed using FINEMAP v1.4 ^34^ on a ±500 kb region centred on the lead SNP, with linkage disequilibrium (LD) estimated from the twin genotype data (one individual per family) and up to 10 causal variants allowed. SNP-based heritability was estimated using GCTA-GREML. GWAS summary statistics were annotated using FUMA v1.6,^35^ including independent significance defined at r^2^ < 0.6 and lead SNPs at r^2^ < 0.1 based of 1000 Genomes Phase 3 (EUR) reference panel, with genomic risk loci delineated by merging LD blocks within 250 kb. Positional mapping, eQTL integration (GTExV8, DICE, PysychENCODE, Fairfax 2014) and functional annotation performed within FUMA. and cross-trait analysis using publicly available summary statistics from FinnGen and the NHGRI-EBI GWAS Catalog.^36^ Methylation quantitative trait locus (mQTL) annotation used in-silico look-up of fine-mapped variants in mQTLdb.^37^

Epigenome-, proteome- and metabolome-wide associations were examined using generalised estimating equations (GEE) implemented in the *geepack* R package,^38^ with CpG sites, proteins and metabolites as outcomes and SLE scores (recent, proximal, lifetime) as predictors. Models were adjusted for age, sex, BMI and smoking status at blood sampling; epigenomic analyses additionally adjusted for methylation bead-chip row. Non-independence of twin pairs was accounted for by specifying an exchangeable correlation structure. Nominal *p*-values were corrected for multiple testing using the Benjamini-Hochberg false discovery rate (FDR) procedure; associations with FDR-corrected *p* < 0.05 were considered statistically significant. Functional annotation of proteomic associations used WebGestalt (www.webgestalt.org) with gene set enrichment analysis (GSEA) based on Gene Ontology Biological Process (GOBP) and Reactome gene sets; all quantified proteins were ranked by signed test statistic. Pathways with FDR < 0.05 were considered statistically significant. All analyses were performed in R (version ≥4.0).

## Results

### Study design and cohort characteristics

The study was conducted within the Older Finnish Twin Cohort, a population-based cohort with longitudinal follow-up from 1975 (Figure 1). Cohort characteristics and layer-specific sample sizes are detailed in Table 1. Across the circulating-omic analyses (epigenomics, proteomics, metabolomics), participants were aged 56–70 years at the time of blood sampling (mean ∼62 years) and predominantly female (∼60%). Mean lifetime SLE scores ranged from 163 to 175 across analytical subsamples; average recent and proximal SLE scores were 65 and 84, respectively, in subsamples with data for those windows.

**Fig 1:**
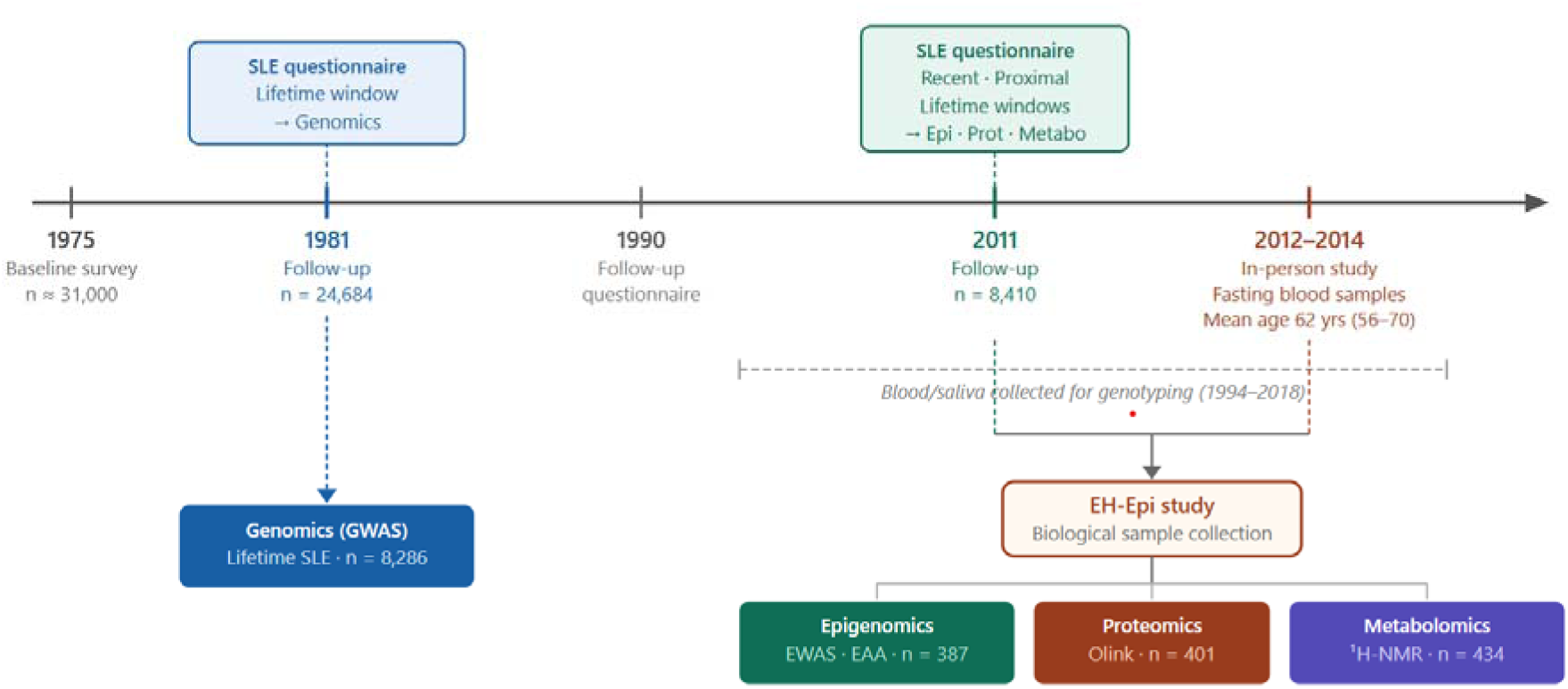
Study design. Genomic analyses used the 1981 SLE assessment, with blood and saliva samples used for genotyping collected between 1994 and 2018. Epigenomic, proteomic and metabolomic analyses used the 2011 SLE assessment, with blood fasting blood samples collected during the EH-Epi in-person styudy (2012-2014). Sample sizes reflect participants with complete data for each molecular layer.SLE, stressful life event; EWAS, epigenome-wide association study; EAA, epigenetic age acceleration; ¹H-NMR, proton nuclear magnetic resonance. Sample sizes reflect participants with complete data for each molecular layer.

### Genome-wide association analysis and heritability

A genome-wide association analysis of lifetime SLE scores in 8 286 individuals identified a single genome-wide significant locus on chromosome 1 (chr1:58 389 816–58 488 601), defined by lead SNP rs10158287 (chr1:58404241:C:T, *p* = 9.7 × 10^-9^) (Figure 2). The locus comprised 118 variants in LD, 95 of which passed the genome-wide significance threshold. After LD pruning, rs10158287 was the sole independent significant SNP. Positional mapping assigned the locus to *DAB1* (Disabled-1), which overlapped all 118 variants. Integration with eQTL data revealed that variants in LD with rs10158287 were associated with downregulated *DAB1* expression specifically in Th1/Th17 CD4^+^ T cells (DICE/T_CD4_TH1_17, *p* = 2.3 × 10^-5^, FDR = 0.049). Additional variants were associated with reduced expression of *C8B* (complement component 8 beta subunit) in naive monocytes (Fairfax_2014_naive, *p* = 6.9 × 10^-7^, FDR = 0.003) and in brain tissue (PsychENCODE, *p* = 6.7 × 10^-5^, FDR = 0.006).

**Fig 2:**
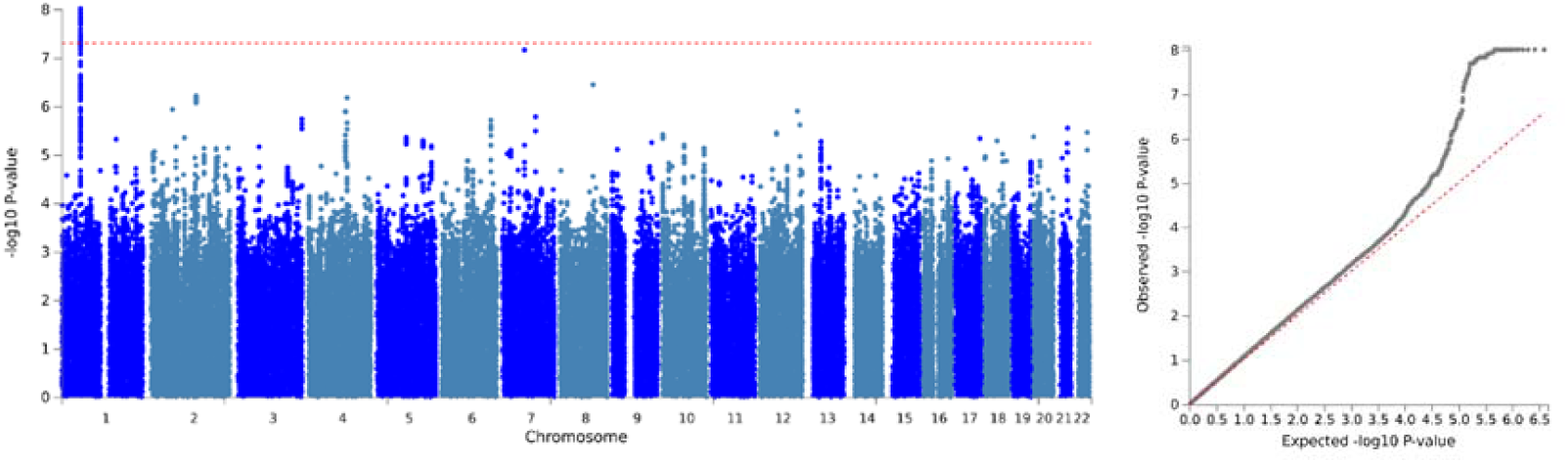
Chromosome 1 locus associated with lifetime stressful events. Manhattan plot (left) of chromosomal position (x-axis) against -log_10_(p) for the association between genetic variants and the SLE score. The dashed horizontal line indicated the genome-wide significance threshold (p = 5 x 10^-8^). The QQ plot (right) shows the distribution of observed versus expected -log_10_(p) values.

Bayesian fine-mapping prioritised a single signal at the chromosome 1 locus, with support concentrated within 58 402 260–58 404 693 bp. Model probabilities favoured a single causal variant (probability = 0.748), although strong LD prevented resolution of a unique causal SNP; the five highest-priority variants (rs10158287, rs11577195, rs10157600, rs1857380, rs1857379) showed comparable posterior inclusion probabilities (PIP = 0.034). Regional SNP-heritability attributable to this locus was estimated at 0.00376 (95% CI 0.00158–0.00660), corresponding to approximately 0.4% of the total SNP-based heritability. mQTL analysis using mQTLdb demonstrated that all five fine-mapped variants were associated with reduced methylation at cg07026918, in *cis* with the *DAB1* locus, at both birth (*p* = 1.40 × 10^-9^) and in childhood (*p* = 1.87 × 10^-9^) (Supplementary Table 1).

Cross-trait analyses in FinnGen identified additional associations at the *DAB1*/*C8B* locus with lipoprotein disorders (*p* = 1.1 × 10^-8^), statin use (*p* = 4.1 × 10^-47^), body mass index (*p* = 7.1 × 10^-9^) and hypertension (*p* = 2.5 × 10^-9^). SNP-based heritability estimated using GCTA-GREML was 33.6% (h^2^ = 0.336, SE = 0.026, *p* = 6.3 × 10^-44^). Twin-based analyses in 425 monozygotic (MZ) and 563 dizygotic (DZ) pairs showed greater within-pair similarity in lifetime SLE scores among MZ than DZ twins (r*MZ* = 0.328, 95% CI 0.24–0.41; r*DZ* = 0.202, 95% CI 0.12–0.28), consistent with a heritable component to SLE exposure.

### Epigenome-wide association study and epigenetic ageing

EWAS was conducted for proximal and lifetime SLE exposure (*n* = 387). EWAS of recent SLE exposure was not performed owing to insufficient sample size within this window for adequately powered testing (*n* = 61 with recent events). No CpG sites were associated with lifetime or proximal SLE exposure after FDR correction. Nominally significant probes are listed in Supplementary Tables 2 and 3.

EAA was assessed across all exposure windows using PCPhenoAge, DunedinPACE and PCGrimAge. No significant associations were observed between SLE scores and any epigenetic ageing measure in the recent, proximal or lifetime windows after correction for multiple testing. All association statistics are provided in Supplementary Table 4.

### Proteomic associations with stressful life events

To investigate systemic signatures of SLEs, we examined associations with 2 321 plasma proteins in 401 participants, stratified by exposure window (Table 2, Figure 3).

**Fig 3:**
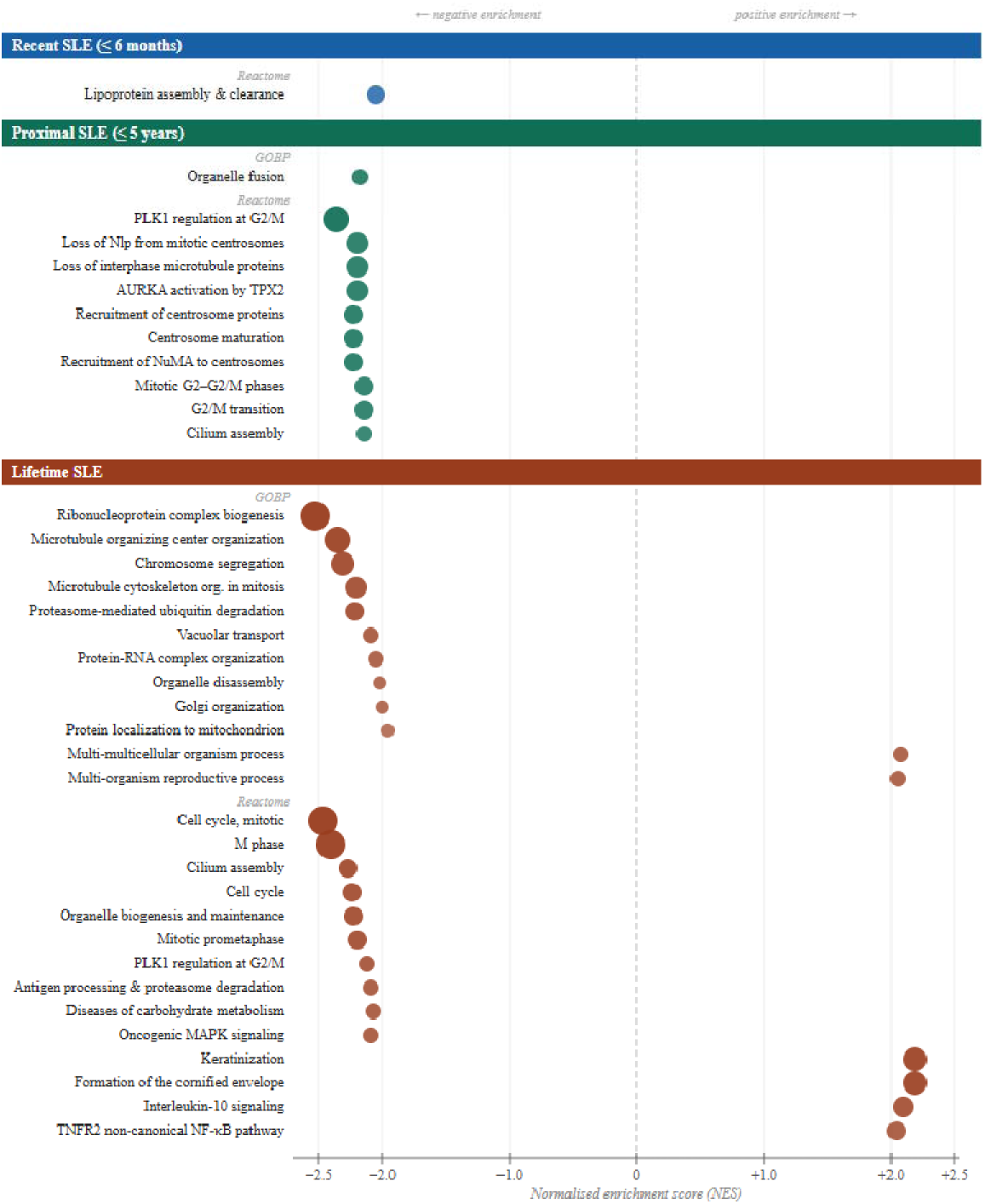
Gene set enrichment analysis of plasma proteomic associations with stressful life events across exposure windows. Gene set enrichment analysis was performed on proteins ranked by signed test statistic. Reactome and Gene Ontology Biological Process databases. All gene sets with FDR < 0.05 are shown. Dot size is proportional to −log□□(FDR); dot position on the x-axis reflects the normalised enrichment score (NES). Colours denote exposure window: blue, recent (≤6 months); teal, proximal (≤5 years); coral, lifetime. FDR, Benjamini–Hochberg false discovery rate; NES, normalised enrichment score.

**Table 2.**
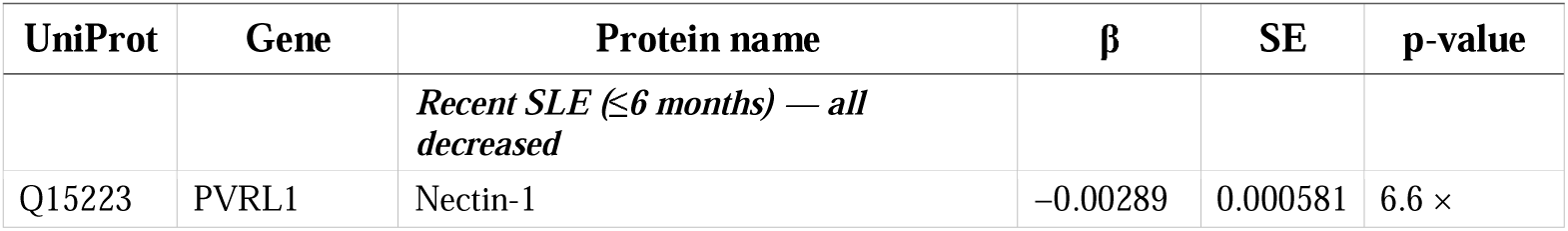
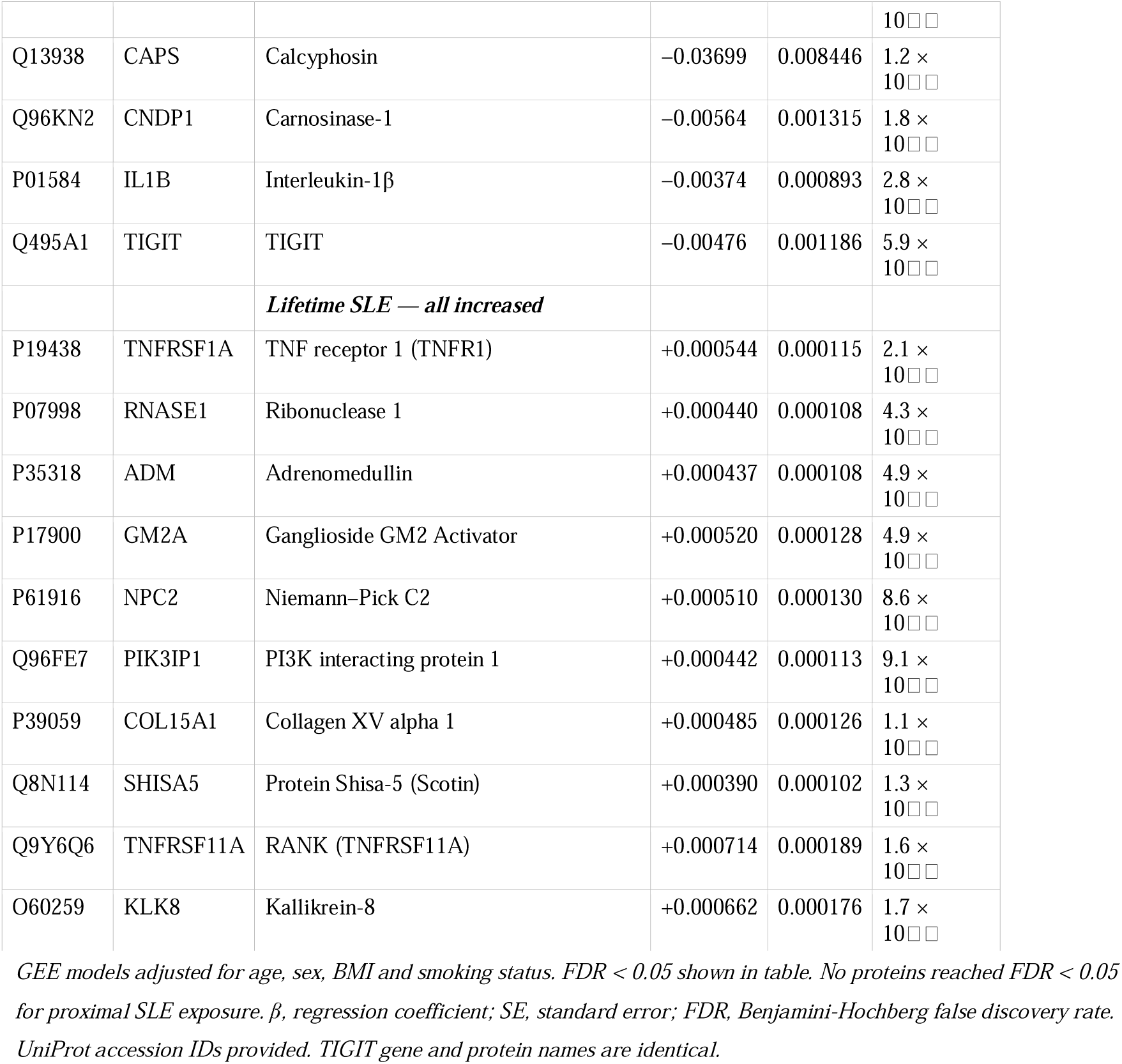
Plasma proteins significantly associated with stressful life events (FDR < 0.05)

#### Recent SLE exposure (≤6 months)

Five proteins were significantly associated with recent SLE burden after FDR correction (FDR < 0.05), all showing reduced circulating levels: Nectin-1 (*PVRL1*), Calcyphosin (*CAPS*), Carnosinase-1 (*CNDP1*), Interleukin-1β (*IL1B*) and T cell immunoreceptor with Ig and ITIM domains (*TIGIT*) (Table 2).

At the pathway level, one Reactome pathway showed significant negative enrichment at FDR < 0.05: Plasma lipoprotein assembly, remodeling, and clearance (NES = −2.05, FDR = 0.016) (Figure 3).

#### Proximal SLE exposure (≤5 years)

No individual proteins passed FDR correction for proximal exposure. However, GSEA identified 10 Reactome pathways with significant negative enrichment (FDR < 0.05) in nominally ranked proteins, all clustering around mitotic progression and centrosome-associated processes. Representative pathways include Regulation of PLK1 Activity at G2/M Transition (NES = −2.36, FDR = 0.001) and Mitotic G2–G2/M phases (NES = −2.14, FDR = 0.007). One GOBP term showed significant negative enrichment: organelle fusion (NES = −2.17, FDR = 0.029). All significantly enriched pathways are listed in Figure 3

#### Lifetime SLE exposure

Ten proteins were associated with lifetime SLE burden at FDR < 0.05, all showing increased circulating levels: TNF receptor superfamily member 1A (*TNFRSF1A*), Ribonuclease A family member 1 (*RNASE1*), Adrenomedullin (*ADM*), Ganglioside GM2 Activator (*GM2A*), Niemann–Pick Disease Type C2 (*NPC2*), Phosphoinositide-3-kinase interacting protein 1 (*PIK3IP1*), Collagen type XV alpha 1 chain (*COL15A1*), Protein Shisa-5 (*SHISA5*), TNF receptor superfamily member 11A (*TNFRSF11A*) and Kallikrein-8 (*KLK8*) (table 2). GSEA of ranked lifetime proteomic associations identified 14 Reactome pathways and 12 GOBP terms with significant enrichment, grouping into four functional categories: cell cycle/mitotic processes (negative enrichment), TNF-linked and IL-10 signalling (positive enrichment), protein degradation/antigen processing (negative enrichment) and epithelial differentiation (positive enrichment) (Figure 3).

### Proteomic age clocks

We examined associations between SLE scores and 22 proteomic ageing measures across the three exposure windows (Table 4). For recent SLEs, greater SLE burden was significantly associated with lower thyroid (β = −1.31 × 10^-4^, FDR = 0.011) and lung (β = −3.35 × 10^-3^, FDR = 0.011) proteomic age estimates. No proteomic clock survived FDR correction for proximal SLEs; at nominal significance (raw *p* < 0.05, FDR < 0.25), brain proteomic age and PAC showed positive nominal associations. For lifetime SLEs, greater cumulative exposure was significantly associated with higher arterial proteomic age (β = 1.08 × 10^-3^, FDR = 0.049). At nominal thresholds, positive associations were additionally observed for thyroid proteomic age, PAC, skin and heart clocks, and the Conventional proteomic age estimate (Table 3).

**Table 3.** Associations between SLE exposure and proteomic age estimates across exposure windows.

| Exposure window | Organ/clock | $\beta$ | SE | p-value | FDR |
| --- | --- | --- | --- | --- | --- |
| **Recent ($\leq 6$ m)** |  |  |  |  |  |
| | Thyroid* | $-1.31 \times 10^{-4}$ | $3.83 \times 10^{-4}$ | $6.5 \times 10^{-4}$ | 0.011 |
| | Lung* | $-3.35 \times 10^{-3}$ | $1.02 \times 10^{-3}$ | $1.0 \times 10^{-3}$ | 0.011 |
| **Proximal ($\leq 5$ y)** |  |  |  |  |  |
| | Brain | $+1.20 \times 10^{-3}$ | $5.41 \times 10^{-4}$ | 0.026 | 0.217 |
| | PAC | $+1.28 \times 10^{-2}$ | $6.29 \times 10^{-3}$ | 0.043 | 0.217 |
| **Lifetime** |  |  |  |  |  |
| | Artery* | $+1.08 \times 10^{-3}$ | $3.52 \times 10^{-4}$ | 0.002 | 0.049 |
| | Thyroid | $+1.72 \times 10^{-4}$ | $7.07 \times 10^{-4}$ | 0.015 | 0.111 |
| | PAC | $+9.46 \times 10^{-3}$ | $3.99 \times 10^{-3}$ | 0.018 | 0.111 |
| | Skin | $+5.48 \times 10^{-4}$ | $2.36 \times 10^{-4}$ | 0.020 | 0.111 |
| | Conventional | $+6.67 \times 10^{-4}$ | $3.32 \times 10^{-4}$ | 0.045 | 0.144 |
| | Heart | $+5.23 \times 10^{-4}$ | $2.61 \times 10^{-4}$ | 0.045 | 0.144 |

*GEE models adjusted for age, sex, BMI and smoking status. \*FDR < 0.05. $\beta$ , regression coefficient; SE, standard error; FDR, Benjamini-Hochberg false discovery rate. PAC, Phenotypic Age Composite proteomic clock; Conventional, mortality-trained composite proteomic age (Goeminne et al.). Only clocks with raw $p < 0.05$ shown.*

**Table 4.** Metabolites significantly associated with recent stressful life event burden (FDR < 0.05)

| Metabolite | $\beta$ | SE | p-value |
| --- | --- | --- | --- |
| Total lipids in small HDL | -0.00984 | 0.002554 | $1.2 \times 10^{-4}$ |
| Total phospholipids in lipoprotein particles | -0.00922 | 0.002462 | $1.8 \times 10^{-4}$ |
| Phosphatidylcholines | -0.00973 | 0.002717 | $3.4 \times 10^{-4}$ |
| Cholines | -0.00918 | 0.002566 | $3.4 \times 10^{-4}$ |
| Phospholipids in small HDL | -0.01012 | 0.002851 | $3.8 \times 10^{-4}$ |
| Phosphoglycerides | -0.00950 | 0.002725 | $4.9 \times 10^{-4}$ |
| Sphingomyelins | -0.00792 | 0.002296 | $5.6 \times 10^{-4}$ |
| Total esterified cholesterol | -0.00801 | 0.002389 | $8.0 \times 10^{-4}$ |
| Concentration of small HDL particles | -0.00766 | 0.002384 | $1.3 \times 10^{-3}$ |
| Glucose | -0.00857 | 0.002743 | $1.8 \times 10^{-3}$ |
| Ratio of saturated FA to total FA | -0.00990 | 0.003169 | $1.8 \times 10^{-3}$ |
| Free cholesterol in small HDL | -0.00966 | 0.003097 | $1.8 \times 10^{-3}$ |
| Saturated fatty acids | -0.01013 | 0.003253 | $1.8 \times 10^{-3}$ |
| Total cholesterol | -0.00807 | 0.002596 | $1.9 \times 10^{-3}$ |
| Total lipids in lipoprotein particles | -0.00933 | 0.003103 | $2.6 \times 10^{-3}$ |
| Total fatty acids | -0.00941 | 0.003213 | $3.4 \times 10^{-3}$ |

*GEE models adjusted for age, sex, BMI and smoking status (n = 434). $\beta$ , regression coefficient (per unit increase in recent SLE score); SE, standard error; FDR, Benjamini-Hochberg false discovery rate. FA, fatty acid; HDL, high-density lipoprotein. All association FDR < 0.05.*

### Metabolomic associations with stressful life events

In metabolomic analyses (*n* = 434), recent SLE exposure was negatively associated with 16 metabolites after FDR correction (Table 4). Associations were predominantly driven by HDL-related lipid measures, including small HDL lipid content, phospholipids and free cholesterol, alongside broader lipid pools (total phospholipids, phosphatidylcholines, sphingomyelins, total and saturated fatty acids, total cholesterol, cholesterol esters) and glucose. No metabolites survived FDR correction for proximal or lifetime SLE exposure.

## Discussion

This study provides an integrated multi-omic view of how SLEs, a partly heritable exposure, are biologically embedded across genomic, epigenomic, proteomic and metabolomic layers. By contrasting three exposure windows, we demonstrate that recent SLEs were primarily associated with coordinated shifts in circulating proteins and metabolites linked to immune regulation and lipoprotein biology, whereas lifetime stress was associated with more stable alterations involving inflammatory signalling, vascular biology and biological ageing, with proximal SLEs showing attenuated single-analyte signals but coherent pathway-level changes in cell-cycle and centrosomal proteins. Across all three time windows and multiple molecular layers, elements of adaptive T cell-related immune regulation recur in different contexts.

At the genetic level, we identified a genome-wide significant locus on chromosome 1 indexed by rs10158287 and mapping to DAB1 and C8B. eQTL analyses demonstrated that variants in LD with the lead SNP were associated with reduced DAB1 expression specifically in Th1/Th17-polarised CD4+ T cells — activated T helper states sensitive to glucocorticoid regulation and linked to chronic inflammatory and immune-mediated disorders^39,40^— and reduced C8B expression in monocytes and brain tissue. While DAB1 is classically characterised for its role in neuronal migration through Reelin signalling,^41^ the cell-type specificity of the regulatory effects observed here implicates immune pathways. C8B encodes a component of the terminal complement complex involved in membrane-attack complex formation,^42^ with complement activity implicated in microglial activation and synaptic remodelling.^43,44^ mQTL annotation showed that these fine-mapped variants are associated with reduced methylation at cg07026918 within the DAB1 locus from birth through childhood, placing the GWAS signal within a genetically anchored regulatory context that precedes adult SLE exposure. This inherited regulatory variation is distinct from experience-dependent methylation changes and does not imply that the GWAS locus mediates adult stress effects on DNA methylation. SNP-based heritability was 33.6%, consistent with a largely polygenic architecture, with the single DAB1/C8B locus accounting for only ∼0.4% of this heritability. Twin-based analyses in the same cohort yielded within-pair correlations of rMZ = 0.328 and rDZ = 0.202, indicating a heritable component to SLE exposure; the convergence between twin-based and SNP-based estimates suggests that additive effects of common variants account for a substantial portion of this heritable signal.^9^ Cross-trait associations with lipoprotein disorders, BMI, hypertension and statin use within the Finnish population indicate cardiometabolic pleiotropy at this locus, consistent with established interconnections between lipid metabolism and immune activation.^45,46^ Cross-layer mediation between the genetic signal and downstream circulating molecular associations cannot be inferred from these data and will require larger longitudinal multi-omic designs.

In contrast to the proteomic and metabolomic findings, no epigenome-wide significant associations were detected for SLEs in whole blood, and no associations with epigenetic ageing measures were observed. The absence of detectable EWAS signals is consistent with the modest sample size (n = 387) and with evidence that DNA methylation effects of common adult psychosocial stressors in peripheral blood may be small and context-dependent,□□ and findings on stress and epigenetic ageing are not consistent across cohorts or stress definitions.□□ Cumulative life stress has been associated with accelerated epigenetic ageing in some population-based samples, with evidence implicating glucocorticoid-mediated regulation,¹□ but our results do not support strong whole-blood methylation associations or epigenetic-ageing acceleration with adult SLEs in this cohort, nothing that tissue-specific effects cannot be excluded. These findings shift interpretive weight toward circulating molecular layers.

The proteomic and metabolomic findings, by contrast, showed that recent and lifetime SLE burden is associated with detectable circulating molecular differences, suggesting that stress-related biological embedding may be captured more readily in time-sensitive circulating phenotypes. For recent SLEs, the five FDR-significant proteins provide convergent context for immune regulation. IL-1β is a central innate cytokine and driver of Th17 programming,^49^ linking the recent-window proteomic profile to the Th1/Th17 immune axis implicated by the DAB1 eQTL, although our data cannot establish whether the genetic signal and the circulating IL-1β association reflect a shared mechanism. Pro-inflammatory cytokines such as IL-1β can impair glucocorticoid receptor signalling,□□ while the Th17 phenotype has been linked to reduced glucocorticoid responsiveness in chronic inflammatory conditions.^51^ TIGIT is an inhibitory receptor expressed on activated T cells and NK cells that regulates effector responses.^52^ The remaining significant proteins, nectin-1 (cell adhesion^53^), Carnosinase-1 (carnosine metabolism and redox pathways^54^) and Calcyphosin (calcium-binding signalling^55^), situate recent SLEs at multiple cellular interfaces. At the pathway level, the most robust signal was negative enrichment of lipoprotein assembly and clearance pathways, accompanied by suppressed HDL-related lipid measures in the metabolomic data. This convergence across proteomics and metabolomics suggests coordinated regulation of lipid metabolism in response to recent stress, consistent with reports linking psychosocial stress to elevated cholesterol^56^ and with stroke risk^57^ and aligns with the cardiometabolic cross-trait annotation at the DAB1/C8B locus. Psychosocial stress is increasingly recognised as engaging central-peripheral neuroimmune pathways that influence inflammatory, lipid and cardiovascular regulation,^58^ and lipid metabolism and immune function are tightly interconnected such that disturbances in cholesterol handling can shape immune activation and inflammatory signalling.^45,46^

For proximal SLEs, the absence of significant single-analyte associations alongside coherent negative pathway enrichment centred on cell-cycle regulation and centrosomal organisation is consistent with relative suppression of reprioritisation of proteins mapping to proliferative and cytoskeletal organisation programmes, without evidence of overt changes in cell division rates.^59,60^

For lifetime SLEs, the ten FDR-significant proteins converge on immune-vascular and tissue remodelling biology. TNFRSF1A (TNFR1), the canonical receptor for TNF-α, is a circulating marker associated with cardiovascular events and mortality;^61^ TNF signalling contributes mechanistically to endothelial dysfunction and arterial stiffening.^62,63^ TNFRSF11A (RANK) participates in NF-κB signalling and has roles in immune activation, lymph node development and bone remodelling,^64^ and has been implicated in multiple cancers.^65,66^ PIK3IP1, a negative regulator of PI3K signalling, modulates T cell activation thresholds and immune homeostasis,^67^ suggesting potential compensatory immune regulation in the context of cumulative stress exposure. Adrenomedullin (ADM) is a vasodilatory peptide induced by inflammatory and haemodynamic stress and widely studied in cardiovascular disease contexts.^68^ RNASE1 is an endothelial-derived ribonuclease involved in maintaining vascular homeostasis and protecting against extracellular RNA-mediated inflammation,^69^ with implications for anti-tumour activity through enhanced T cell activation,^70^ though its dysregulation during prolonged inflammation is associated with vascular disease^69^. COL15A1 encodes a basement membrane-associated collagen implicated in vascular integrity and remodelling.^71^ GM2A and NPC2 are lysosomal lipid-transport proteins central to sphingolipid and cholesterol handling.^72,73^ SHISA5 is a p53-inducible protein involved in apoptosis and cellular stress responses through NF-κB regulation,^74,75^ while KLK8 participates in extracellular proteolysis with roles in neural plasticity and implications for psychiatric disorders,^76,77^ skin inflammation^78^ and several cancers.^79,80,81^ At the pathway level, the bidirectionality of lifetime enrichment — simultaneous positive enrichment of TNF/IL-10 signalling and epithelial differentiation alongside negative enrichment of cell-cycle programmes — suggests concurrent inflammatory activation and regulatory modulation within cytokine networks. IL-10 signalling provides immunoregulatory counterbalance within inflammatory systems,^82^ and its positive enrichment alongside TNF programmes is consistent with a consolidated immune-vascular phenotype. Epithelial differentiation ontologies suggest that cumulative SLE burden is reflected in circulating proteins mapping to skin-associated structural programmes, consistent with evidence that stress alters epidermal barrier regulation,^83^ though plasma proteomics does not allow tissue-specific attribution.

Metabolomic profiling further supported a time-dependent signature aligned with the recent-window lipoprotein pathway signal. Only recent SLE exposure showed FDR-significant metabolites, all negative: multiple HDL-related measures alongside broader lipid pools and glucose. These results are consistent with the concept of lipid reactivity to psychosocial stress observed in controlled paradigms, although the direction and magnitude of lipid changes can vary with context and population.^84^ No associations survived corrections at proximal and lifetime windows.

We observed exposure-window differences in proteomic ageing measures. In the recent window, greater SLE burden was associated with lower thyroid and lung proteomic age estimates, whereas in the lifetime window the arterial proteomic age estimate was significantly higher, converging with the lifetime proteomic profile (TNFRSF1A, ADM, RNASE1, COL15A1) and TNF-linked pathway enrichment, consistent with vascular remodelling and inflammatory ageing biology.^85,86^ This directional shift, from lower organ-specific age estimates in the recent window to higher arterial age in the lifetime window, is consistent with a transition from short-term physiological adaptation to longer-term alignment with cumulative exposure. In the proximal window, no clocks survived FDR correction, but nominal positive associations were observed for brain age and PAC. At nominal thresholds in the lifetime window, PAC and conventional proteomic age, which reflect aggregated multisystem ageing patterns rather than single-organ processes^86^, alongside heart and skin clocks were also positively associated, concordant with the vascular and epithelial signatures noted above.

Across layers, the associations with SLE exposure did not simply intensify over time; they shifted in emphasis. Rather than linear accumulation, the data support temporal redistribution of molecular signatures: from acute immune-metabolic reactivity (recent) through pathway-level reprogramming (proximal) to consolidated immune-vascular signatures (lifetime). This pattern unfolds against a partly heritable variation in SLE exposure, largely accounted for by common variants. The DAB1/C8B locus provides a genomically anchored immune-regulatory signal that remains directionally consistent with downstream immune themes across windows, without implying direct causal mediation, which will require larger longitudinal multi-omic designs with mediation and path-analytic approaches in genetically informative samples.

### Strengths and Limitations

This study has several important strengths. First, it leverages a prospectively followed population-based twin cohort with longitudinal data spanning four decades, enabling assessment of SLE exposure across multiple life-course windows with robust temporal framing. Second, the SLE measure employs the Holmes and Rahe Social Readjustment Rating Scale,^12^ whose individual components have been independently linked to clinically meaningful endpoints including cardiovascular mortality and cancer risk.^6,14^ Third, the concurrent availability of four omic layers (genome, epigenome, proteome, metabolome) from the same cohort enables cross-layer triangulation of findings in a manner that single-omic studies cannot. Fourth, the twin design enables partitioning of genetic and environmental contributions to stress exposure and to molecular variability, providing a unique resource for genetically informed inference.

A few limitations warrant consideration, nonetheless. First, sample sizes for the circulating omic layers (*n* = 387–434) are modest relative to current standards for EWAS and proteomic analyses, limiting power to detect small effects. The absence of significant EWAS signals may partly reflect this constraint. Second, the cohort consists exclusively of individuals of Finnish ancestry, which, while conferring genetic homogeneity and reducing population stratification, limits generalisability to more diverse populations. Third, there is a temporal gap between SLE questionnaire assessment (2011) and biological sample collection (2012–2014; mean gap ∼2 years), which may have attenuated associations with more transient molecular states, particularly for recent SLE exposure. Finally, the cross-sectional nature of the molecular measurements precludes inference about the temporal ordering of molecular changes, and longitudinal within-person molecular data will be required to establish causal directionality; longitudinal designs with mediation approaches in genetically informative samples represent important next steps.

## Conclusion

The present findings demonstrate that the molecular embedding of stressful life events is temporally structed rather than uniform, with genomic, proteomic and metabolomic signatures shifting in biological emphasis across recent, proximal and lifetime windows. This temporal patterning, spanning lipid-immune reactivity, pathway-level cytoskeletal reprogramming and consolidated immune-vascular remodelling, argues against a simple accumulation model of stress biology and points instead to window-specific molecular responses. Across this progression, adaptive T cell-related immune regulation emerges as a recurrent theme at multiple layers of biological organisation, from genetically anchored regulatory variation at the DAB1/C8B locus to circulating inflammatory and vascular proteins and enriched cytokine signalling pathways. Whether these molecular signatures lie on causal pathways to stress-related disease outcomes remains an important open question, and larger prospective multi-omic studies will be well placed to address it.

## Data Availability

Data from the Finnish Twin Cohort are not publicly available due to restrictions of informed consent. Data are available to authorised researchers through the Institute for Molecular Medicine Finland (FIMM) Data Access Committee (DAC), subject to IRB/ethics approval, an institutionally approved study plan, and a data use/transfer agreement in compliance with national data protection legislation. Summary statistics from the proteomic and metabolomic analyses are available from the corresponding author upon reasonable request.

